# Scenario Design for Infectious Disease Projections: Integrating Concepts from Decision Analysis and Experimental Design

**DOI:** 10.1101/2023.10.11.23296887

**Authors:** Michael C. Runge, Katriona Shea, Emily Howerton, Katie Yan, Harry Hochheiser, Erik Rosenstrom, William J.M. Probert, Rebecca Borchering, Madhav V. Marathe, Bryan Lewis, Srinivasan Venkatramanan, Shaun Truelove, Justin Lessler, Cécile Viboud

## Abstract

Across many fields, scenario modeling has become an important tool for exploring long-term projections and how they might depend on potential interventions and critical uncertainties, with relevance to both decision makers and scientists. In the past decade, and especially during the COVID-19 pandemic, the field of epidemiology has seen substantial growth in the use of scenario projections. Multiple scenarios are often projected at the same time, allowing important comparisons that can guide the choice of intervention, the prioritization of research topics, or public communication. The design of the scenarios is central to their ability to inform important questions. In this paper, we draw on the fields of decision analysis and statistical design of experiments to propose a framework for scenario design in epidemiology, with relevance also to other fields. We identify six different fundamental purposes for scenario designs (decision making, sensitivity analysis, value of information, situational awareness, horizon scanning, and forecasting) and discuss how those purposes guide the structure of scenarios. We discuss other aspects of the content and process of scenario design, broadly for all settings and specifically for multi-model ensemble projections. As an illustrative case study, we examine the first 17 rounds of scenarios from the U.S. COVID-19 Scenario Modeling Hub, then reflect on future advancements that could improve the design of scenarios in epidemiological settings.

## Introduction

Epidemics prompt many questions, from public health policy makers wanting to know how to intervene, to members of the public wanting to know what to expect, to industrial leaders wanting to know how to react. These questions are all necessarily forward looking, creating a demand for epidemiologists to project what may happen in the future. Epidemiological models provide a way to integrate historical observations, biological and sociological knowledge, and our understanding of disease mechanisms to produce projections of epidemiological outcomes into the future. These projections can be used to guide decisions (by governments, industries, and individuals) about how to respond, and to guide research investment (to reduce uncertainty in projections or processes).

Quantitative scientists make a distinction between forecasts and scenario projections (Reich et al., 2022). Forecasts are unconditional predictions about the future, statements about what is expected to happen. The most useful forecasts are probabilistic, expressly recognizing and transparently quantifying the uncertainty in the prediction. Scenario projections, on the other hand, are conditional predictions about the future, statements about what would happen if a set of conditions were to be met. The most useful scenario projections would also be probabilistic, but conditionally so; they typically express the probability of outcomes if certain conditions are met, but do not usually quantify the probability of those conditions themselves being met. Scenario projections, then, are exploratory—they allow the examination and contrast of multiple futures.

Scenario modeling is common in many disciplines, including climate science (Krey, 2014), conservation biology (Nicholson et al., 2019), wildlife and fisheries management (Johnson et al., 1997), economics (McDowall and Eames, 2006), transportation (Bartholomew, 2007), urban planning (Khakee, 1991), energy development (Leung and Yang, 2012), agriculture (Pfister et al., 2011), invasion ecology (Shea et al., 2005), military planning (Dowse, 2021), disaster planning and response (Tyszkiewicz et al., 2012), nuclear war and terrorism (NASEM, 2023), and many others. One of the most visible global examples is the Coupled Model Intercomparison Project (CMIP), which has produced six phases of climate projections based on shared scenario specifications (Eyring et al., 2016; Meehl et al., 2000). The most recent phase of projections (CMIP6) is based on a set of “shared socio-economic pathways” and provides central evidence for the sixth assessment report of the Intergovernmental Panel on Climate Change. Importantly, the shared socio-economic pathways (and the “representative concentration pathways” of CMIP5) represent forcing scenarios (notably concerning carbon emissions); the climate projections are conditional on the scenario assumptions, but the likelihood of those conditions occurring was not estimated. Similar types of scenario projections are made in many other fields, but there is not yet a common lexicon that unites the large literature on this subject. In an influential book, Martelli (2014) argued that the field of scenario planning faces a number of shortcomings, notably a lack of clarity in the conceptual foundations, methodological inconsistency, and absence of evidence of effectiveness. A more recent review finds progress toward a synthesis of concepts and methods and increasing evidence of effectiveness, but notes that the field remains fragmented (Cordova-Pozo and Rouwette, 2023).

The use of scenario modeling has become pervasive in infectious disease epidemiology over the last two decades. Notable examples include modeling of different types and layers of interventions to control emerging outbreaks such as foot-and-mouth disease in the United Kingdom (Tildesley et al., 2006), avian influenza (Longini et al., 2005), the Ebola outbreak in West Africa (Meltzer et al., 2014), and the COVID-19 pandemic (Borchering et al., 2023; Borchering et al., 2021; Hellewell et al., 2020; Truelove et al., 2022; Walker et al., 2020). Additional notable use cases include the roll-out of new interventions for endemic pathogens where, for instance, scenario projections can help anticipate the benefits and dynamic changes associated with new vaccines or improved drugs (Flasche et al., 2016; Pitzer et al., 2009; Eaton et al., 2012; Houben et al., 2016). In some cases, the scenario projections are produced from a single model (e.g., Meltzer et al., 2014), while in others, the scenario projections come from multiple models (Flasche et al., 2016; Houben et al., 2016), drawing on a growing literature documenting the value of multi-model efforts (Johansson et al., 2019; Shea et al., 2020, 2023; Cramer et al., 2022; Prasad et al., 2023). Scenario design plays an important role in infectious disease projections over long time scales, not only to contrast different intervention schemes, but also to control for uncertainty in key parameters that may be magnified over time. However, there is little guidance on how to optimize scenario assumptions to answer particular public health questions, especially in the context of multi-model efforts.

Across fields, one of the central features of scenario projections is that “scenarios seem to exist in sets and the scenarios that inhabit those sets are systematically prepared to co-exist as meaningfully different alternatives to one another” (Spaniol and Rowland, 2019). How, then, are these sets developed? Many methods for scenario design exist and attempts have been made to classify the methods into several schools of approach (Amer et al., 2013). In this paper, we draw from the fields of decision analysis and experimental design to propose a framework for scenario development that integrates the three schools discussed by Amer et al. (2013). We place this work in the context of epidemiological modeling, but intend the framework to be more broadly useful. Our primary thesis is that clarity about the purpose of the scenarios is central to their design, and we offer a taxonomy of design purposes.

### Purposes of Scenario Design

We approach scenario design like experimental design. First, a scenario design, like an experimental design, should have a purpose—a question (or questions) that the designers seek to answer. Second, a scenario design consists of a set of alternative scenarios (analogous to experimental treatments), which differ with regard to one or a few factors. Third, the foundational experimental design concepts of control, randomization, and replication have analogs in scenario design. The scenarios can be designed to control certain factors by prescribing shared assumptions or parameter values. By randomly sampling from the probability distributions for uncontrolled parameters, inferences from scenario comparisons can be extended to the full parameter space represented by those distributions. Controlling for these otherwise unspecified variables is common in experimental design through a method called pairing; this is also possible in scenario modeling by pairing replicates across scenarios (i.e., compare replicates with the same uncontrolled parameters, that therefore differ only by scenario). Further, each scenario can be replicated many times, either by soliciting repeat projections from a single model structure or by soliciting projections from multiple models of varying structure. These concepts are embedded in our framework for scenario design.

One of the central questions in scenario design is how the individual scenarios differ. Many factors that will affect future dynamics are unknown at the time of projection (e.g., human behavior or key aspects of pathogen biology). Scenario design, then, is the process of strategically choosing among those many uncertainties to identify a set of scenarios that together can achieve the purpose of the scenario projection exercise. Inspired by multiple rounds of COVID-19 projections that have addressed public health goals at different stages of the pandemic, we identify three primary purposes in scenario design: making decisions, exploring uncertainty, and identifying how decisions may be affected by uncertainty. To understand the differences between these purposes, we distinguish two types of factors (often described as “scenario axes”): interventions (or decision options) and uncertainties. Interventions are factors that are under the control (or partial control) of one or more decision makers, such as vaccination policies, non-pharmaceutical intervention (NPI) policies, or hospital staffing and capacity.

Uncertainties are factors that are not under any decision maker’s control, but that might affect the outcomes or possibly even the choice of intervention. This distinction between intervention and uncertainty factors is not always sharp, and can depend on the primary audience. For instance, the arrival of a new virus variant will always be considered uncertain, as its emergence is beyond anyone’s control. However, other factors, such as vaccine coverage, are more complicated, as they can be affected by the informational campaigns of public health agencies (a decision) and the behavioral responses of individuals (an uncertainty). Further, a factor that is an intervention for one decision maker (e.g., vaccination recommendation by the Centers for Disease Control and Prevention, CDC) might be an uncertainty for another decision maker (e.g., a hospital complex). The explicit purpose of a scenario design and its intended primary audience, however, can help shed light on whether a factor should be treated as a decision or an uncertainty.

#### Purposes of Scenario Design: a Taxonomy

Scenario modeling is an attempt to glimpse something about the future, often with the intention of informing actions in the present. In this sense, there is a decision-making element to scenario modeling, but the decisions can have many purposes: to change the trajectory of the future through interventions; to respond to future outcomes; or to seek more information. We believe that understanding the purpose of a scenario modeling exercise informs the design of the scenarios, as well as any subsequent ability to evaluate the success of the exercise. We propose six classes of scenario design that stem from the three primary goals. Two-factor designs are very common in scenario modeling across all fields, so we provide shorthand for each class based on what the two factors would be, but note later that simpler (one factor) or more complex designs are possible.

##### *Decision making* (decision x decision in a 2×2 matrix design)

In a decision-making setting, the scenarios are designed to contrast alternative interventions, actions that are intended to influence the outcomes being modeled. For example, in Figure 1A, the scenario design consists of three scenarios, varying spatial extents of an intervention (nowhere, in 1 of 3 candidate locations, everywhere). If multiple types of interventions are being considered, a factorial arrangement of the levels of each might be of interest, so the scenario design could be, say, a 2×2 matrix with both axes being interventions. The interventions could be alternatives being considered by a decision maker (e.g., Borchering et al., 2023) or potential interventions being explored to nudge decision makers to consider new options (Meltzer et al., 2014).

**Figure 1.**
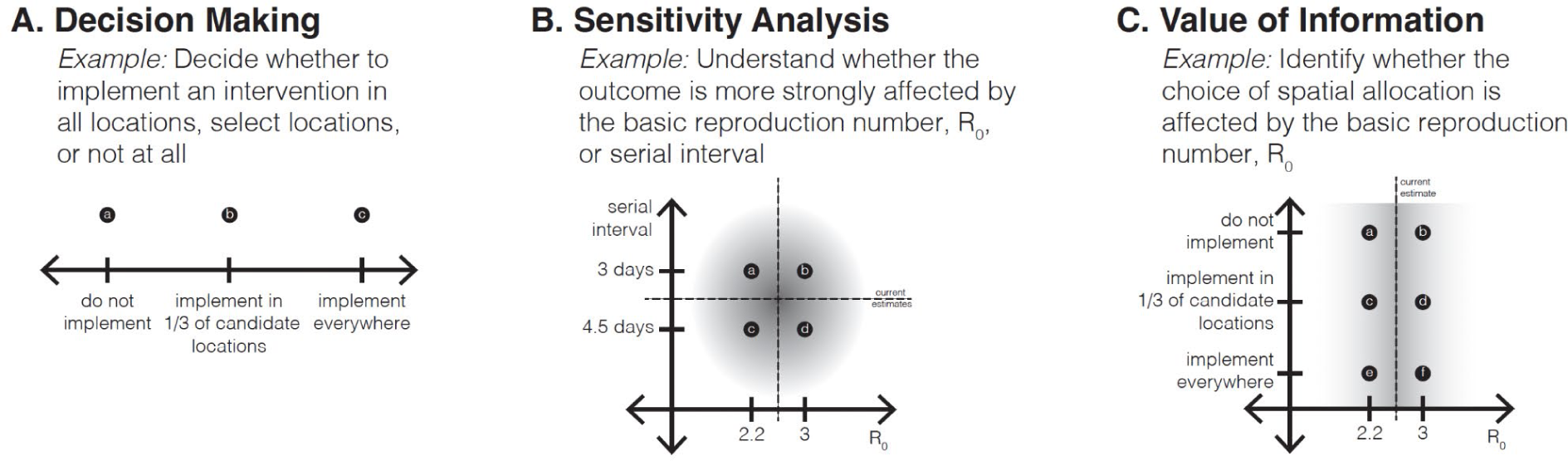
Graphical depiction of three classes of scenario designs, with heuristic examples. (A) In a Decision Making scenario design, the axis or axes are variables that are under the control of the decision maker; the purpose of the design is to understand the outcomes associated with different interventions. (B) Sensitivity Analysis designs focus on understanding the role of different sources of uncertainty on the outcomes of interest. (C) A Value of Information (VOI) design (decision axis x uncertainty axis) examines whether a source of uncertainty affects the relative effects of interventions. The shaded regions represent the current confidence intervals for the uncertainty parameters.

##### *Sensitivity analysis* (uncertainty x uncertainty in a 2×2 matrix design)

The purpose of sensitivity analysis is to understand the contributions of different sources of uncertainty to the outcomes of interest (Saltelli et al., 2004), and potentially whether they interact. In a sensitivity analysis setting, then, the scenario axes focus on uncertainties. For example, in the 2×2 scenario design in Figure 1B, one axis captures uncertainty about the basic reproductive number (*R*0 of 2.2 or 3.0) and the other captures uncertainty about the serial interval (3 or 4.5 days). Note that there is no decision explicit in this design, although research efforts could be devoted to the more influential factor.

##### *Value of information* (decision x uncertainty)

Value of information (VOI) is a common concept in the field of decision analysis that assesses whether the more effective intervention (rather than its outcome) is sensitive to the uncertainty. Thus, value of information is a form of sensitivity analysis from the standpoint of the decision maker (Felli and Hazen, 1998). In a value of information design, at least one scenario axis is an intervention and at least one other is an uncertainty. For example, in Figure 1C, the design consists of 6 scenarios, in a 2×3 design, with one decision axis (the same spatial implementations as in Fig. 1A) and one uncertainty axis (*R*0); the result of particular interest would be whether the ranking of decision options was different under the two values of the basic reproductive number. From a decision-making perspective, this is the most important design, because it evaluates intervention alternatives while also investigating whether their performance is robust to major sources of uncertainty; it also can be used to inform the value of gathering more information.

##### *Situational awareness* (uncertainty x uncertainty)

The remaining three classes of scenario design resemble sensitivity analysis designs, in that the scenario axes focus on uncertainties only, but their purposes differ, possibly affecting construction of the scenarios. Sometimes scenarios are used for situational awareness, to give decision makers and the public a sense of the current state-of-the-world and what might be coming (Fig. 2A). In this way, different from the sensitivity analysis class, decisions are implied, although their effects are not embedded in the design.

**Figure 2.**
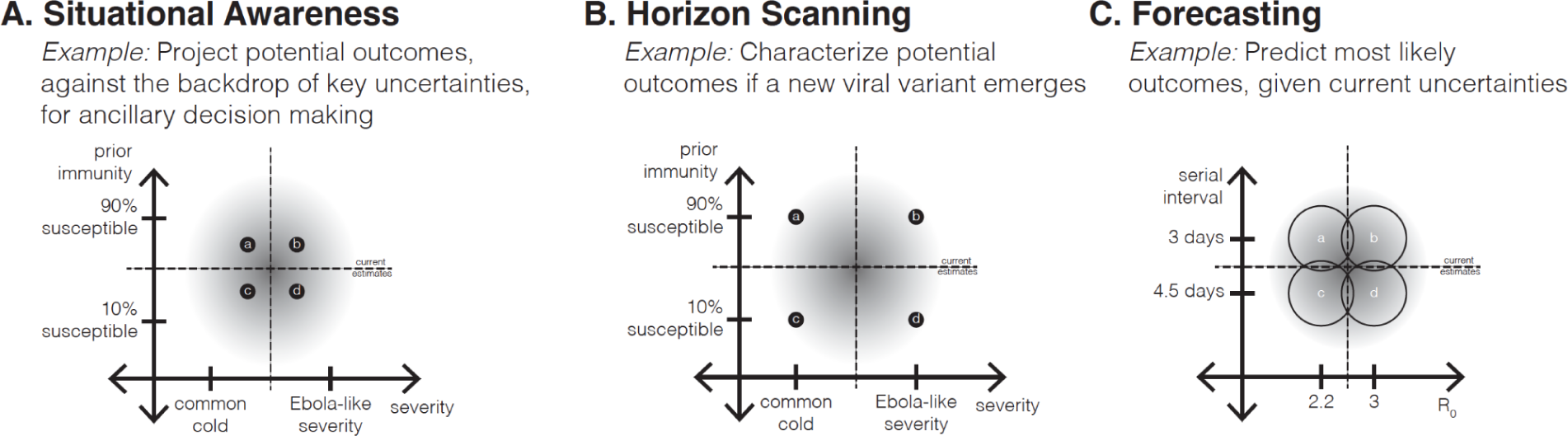
Scenario design classes related to Sensitivity Analysis designs (uncertainty x uncertainty) that have decision-adjacent purposes. (A) Situational Awareness designs may appear indistinguishable from Sensitivity Analysis designs, but have an additional purpose to provide insight about potential outcomes that may be relevant for ancillary decisions. (B) Horizon Scanning designs explore the edges of the epistemic uncertainty, often to prompt insights about what could happen in the future, in an effort to develop new interventions. (C) Forecasting designs postulate multiple hypotheses in the parameter space, with an appropriately weighted average of outcomes constituting a well-calibrated forecast, given the current uncertainty. The shaded regions represent the current confidence intervals for the uncertainty parameters.

##### *Horizon scanning* (uncertainty x uncertainty)

There is a large literature, primarily outside of epidemiology, that focuses on horizon scanning (also often called scenario planning; Sutherland and Woodroof, 2009). In this approach, scenarios are designed to explore plausible extremes of what could happen (Fig. 2B), as a way to provoke awareness of future possibilities, motivate preparation, avoid or plan for surprises, and encourage creation of new intervention strategies. Even though there are no interventions on the axes of this design, this approach is more decision-centric than sensitivity analysis or situational awareness, in that a decision maker is aware of looming threats and is looking for insight to guide novel interventions. In conservation settings, horizon scanning around the possible impacts of climate change has become an important approach, as natural resource management agencies realize that their old tools may no longer be effective in changing ecosystems. In epidemiological settings, the horizon scanning class has been used particularly in thinking about emergence of novel pathogens or variants, like spillover of avian influenza to humans (Colizza et al., 2007).

##### *Forecasting* (uncertainty x uncertainty)

The final approach aims to design a set of scenarios that can be combined into an unconditional probabilistic forecast of the future, by careful choice of scenarios to bracket key uncertainties (Fig. 2C). This approach differs fundamentally from the other types of designs described above: first, the set of scenarios needs to collectively represent the full degree of uncertainty about influential parameters (e.g., those included in analytical expressions for the reproduction numbers derived from mechanistic models); and second, the likelihood of the individual scenarios needs to be specified (or derivable from experience). With these conditions, a weighted combination of the scenario projections forms a proper forecast with appropriate uncertainty. The belief weights on the scenarios (i.e., likelihood of each scenario) can be updated dynamically in time as new evidence comes in using a Bayesian approach or its generalization, Dempster-Shafer Theory (Shafer, 1990). In the field of natural resource management, when such dynamic scenario forecasting is embedded in a Markov decision process, it is called “adaptive management” (Chadès et al., 2012; Walters, 1986); similar approaches are commonly used in machine learning and artificial intelligence applications (Sutton and Barto, 2018).

There are other uses of scenario modeling, in training and tabletop exercises, where the users’ interactions with the scenarios are central to their purpose. The use of scenario modeling has a rich history in military training (Straus et al., 2019; Kim et al. 2014; NRC, 2008), as well as other fields. The goals of tabletop exercises include understanding inter-agency coordination, preparedness in terms of personnel, equipment, and protocols, and other aspects of complex responses to emerging threats. Policy and decision makers are assigned roles and asked to make various decisions during an evolving scenario. Examples of settings where scenario modeling has been used in tabletop exercises include responses to a novel SARS-like agent (Dausey et al., 2005), release of plague bacteria (*Yersinia pestis*, Henderson et al. 2001), and a new outbreak of foot-and-mouth disease in the United Kingdom (DEFRA, 2018). The design of scenarios in these types of exercises tends to be more complex than the others described above, with nested and branching scenarios that respond to user actions. The details of such designs are beyond the scope of this paper, although many of the elements that we discuss will be relevant.

### Scenario Designs Used by the COVID-19 Scenario Modeling Hub

To illustrate the proposed scenario classification, we retrospectively analyzed 17 rounds of scenario designs developed by the U.S. COVID-19 Scenario Modeling Hub (SMH) (https://covid19scenariomodelinghub.org/). Since December 2020, the SMH has convened multiple modeling teams to generate scenario-based projections of COVID-19 cases, hospitalizations, and deaths over 3-24 month horizons, in close collaboration with U.S. public health agencies. The 17 rounds of scenarios available for study addressed different needs at different stages of the pandemic. The scenario classification described in this paper (Figs. 1 and 2) was not available when SMH scenarios were designed, but we have applied it retrospectively, recapturing the intent of each round through publicly released reports and internal notes taken during the design process (Table 1). Scenarios were typically designed through an iterative discussion process between the SMH coordination group, participating modeling teams, and public health partners. This process took anywhere from 3-86 days (median 32.5 days; see also Loo et al., *in review* for more details). For each round, we have identified the “motivating audience” as the decision-making body that the designers had foremost in mind. In most rounds, the motivating audience was the collection of federal, state, and local public health agencies with authority to set public health policy or guidance, but in 6 of the 17 rounds, the design was more strongly motivated by consultation with a specific public health partner (notated in Table 1 with bold type). It is important to note, however, that the SMH coordination group always worked with, and had in mind, the needs of multiple decision-making agencies beyond the motivating audience, and, at times, these considerations also influenced the scenario design.

**Table 1.**
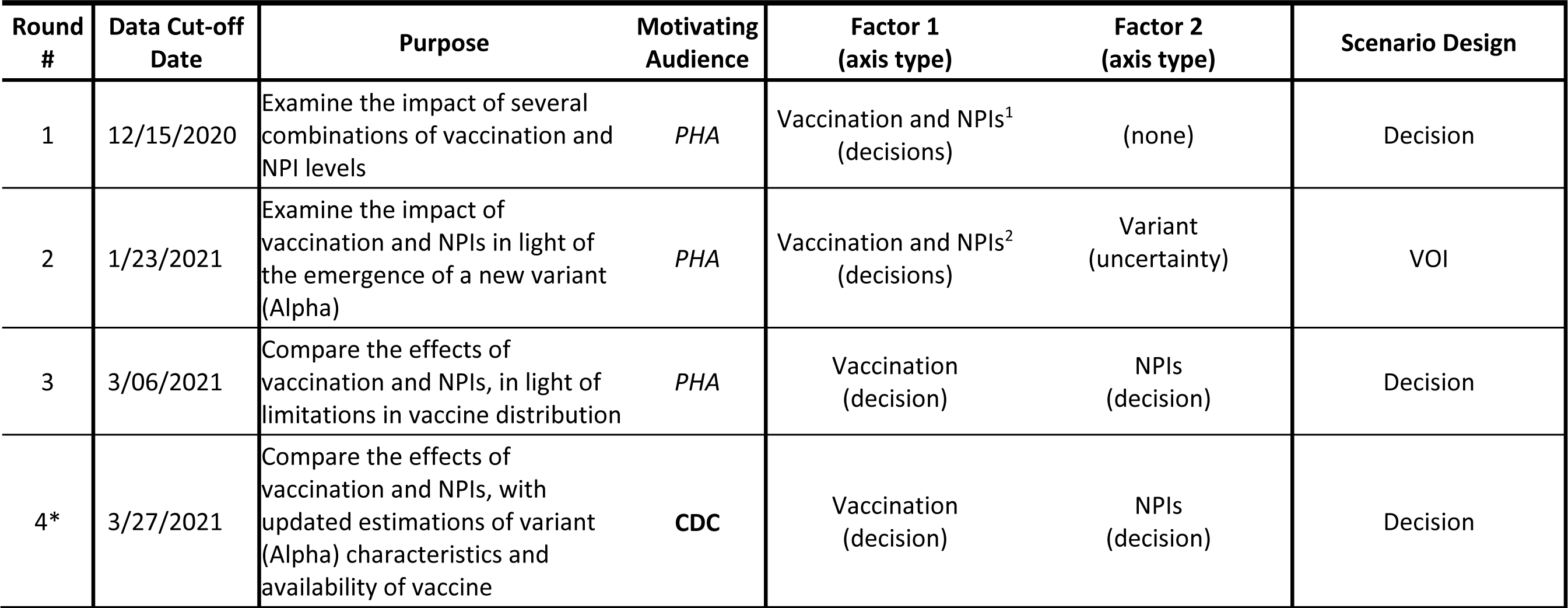
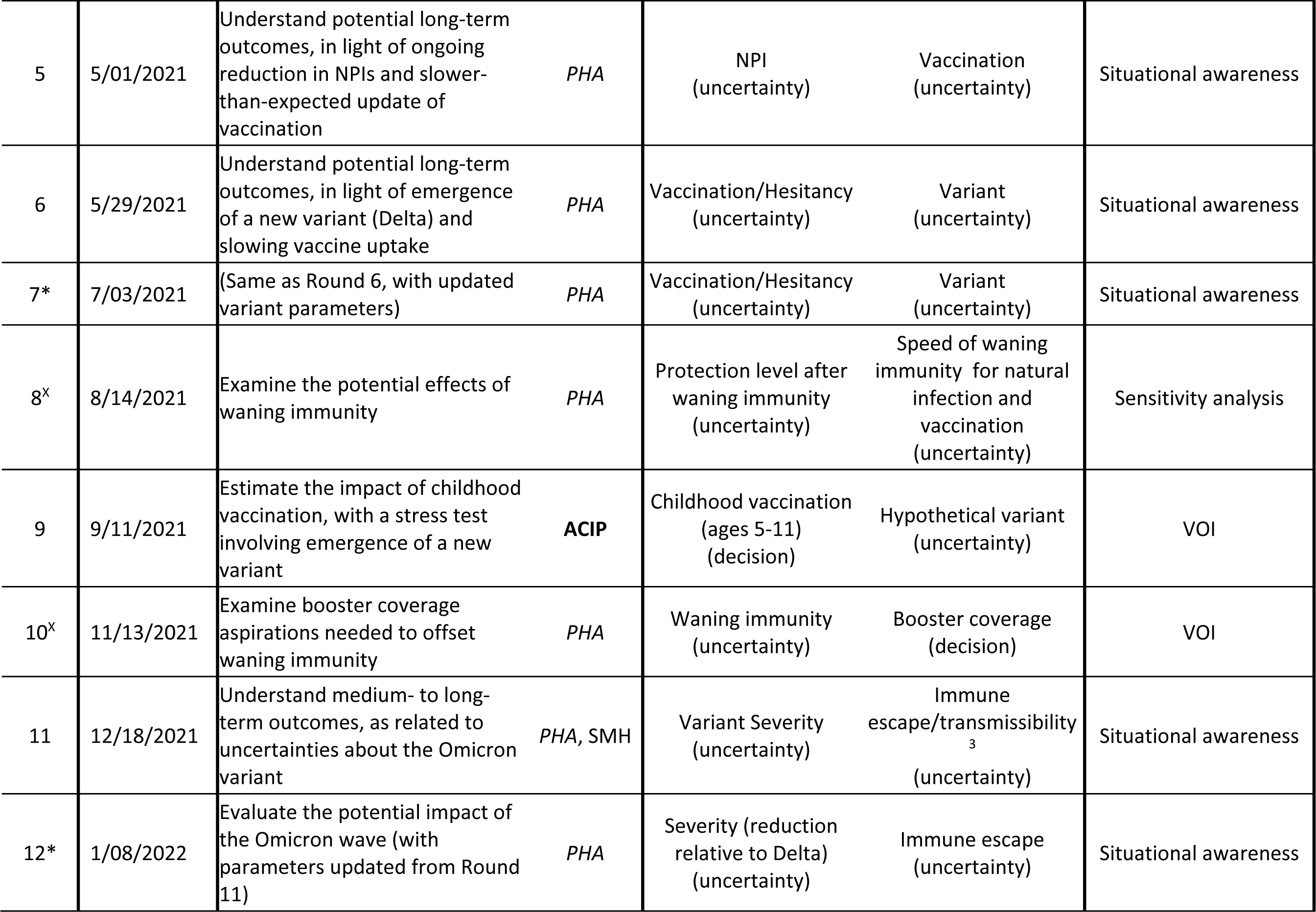
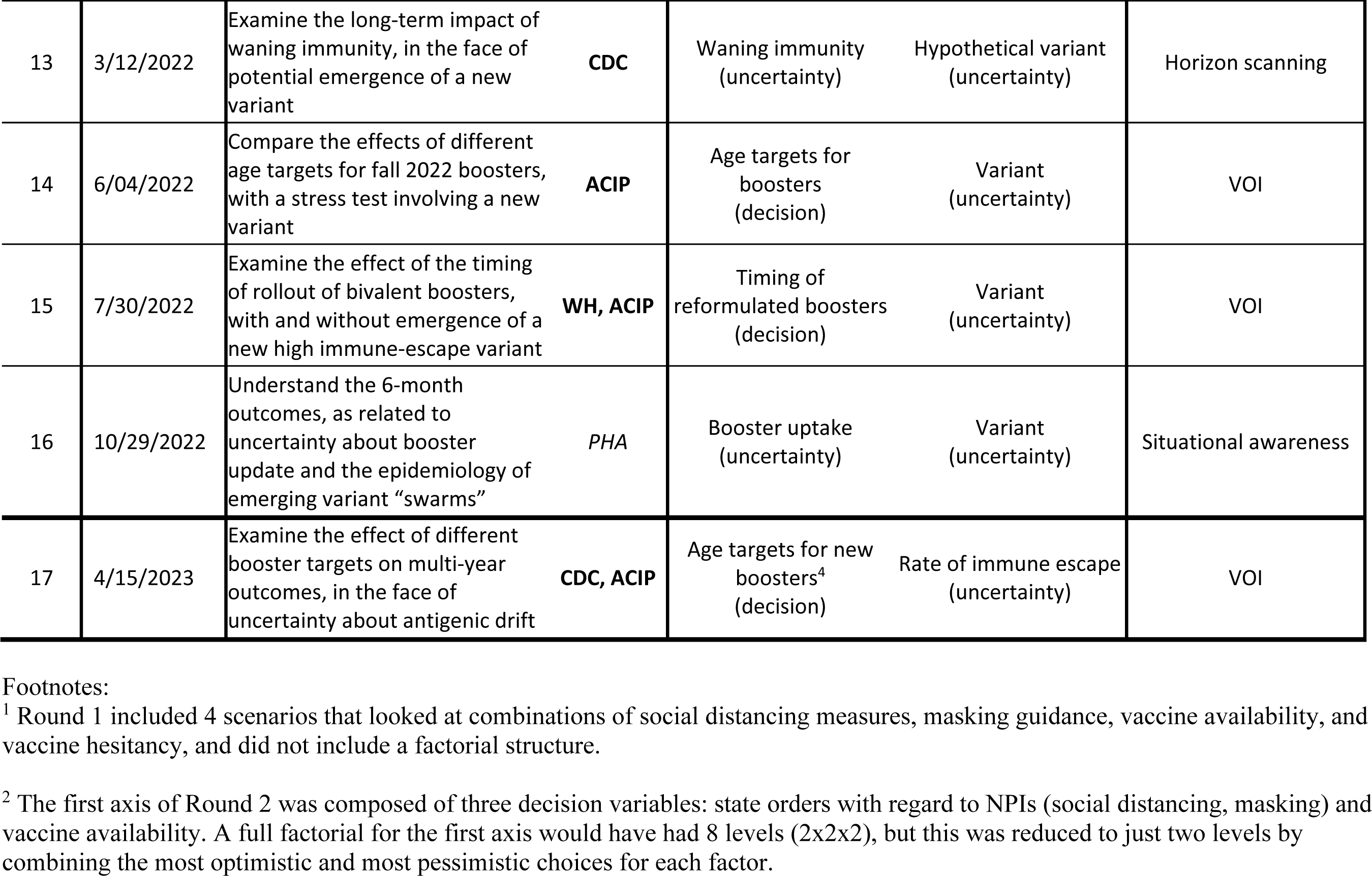
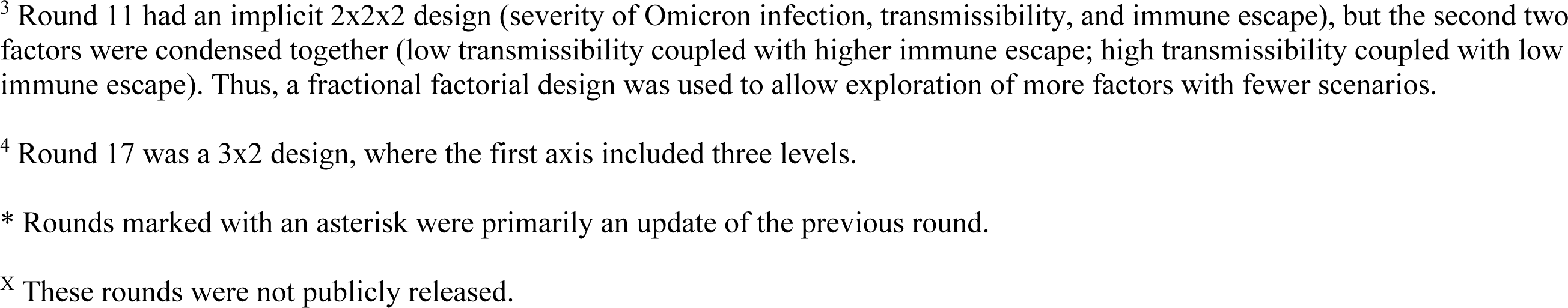
Retrospective determination of scenario designs used by the U.S. COVID-19 Scenario Modeling Hub (SMH) in its first 17 rounds, released February 2021-April 2023. Most SMH rounds included 4 scenarios reflecting different levels of controls or epidemiological situation, depending on the stage of the epidemic. All rounds, except for Rounds 1 and 17, were organized as a 2×2 table representing two axes or key epidemic drivers (e.g., vaccination and NPI, Factors 1 and 2 columns), with a high and low value assumed for each of these drivers. The classification of scenario design (last column) arises from considering the axes types, as well as the purpose of the round: a “Decision” class arises when all scenarios axes are decision axes; a value of information (VOI) class arises when a decision axis is crossed with an uncertainty axis; and the remaining types (sensitivity analysis, situational awareness, and horizon scanning) arise from uncertainty by uncertainty structures. The classification of the axis types was done with reference to the motivating audience, but other audiences could use the results for other purposes. Audience abbreviations: *PHA*, public health agencies, that is, federal, state, or local public health decision makers (*default generic audience* for most rounds); CDC, U.S. Centers for Disease Control and Prevention; WH, the White House COVID-19 Task Force; ACIP, the CDC Advisory Committee on Immunization Practices; SMH, the U.S. COVID-19 Scenario Modeling Hub (for internal insights). Other abbreviations: NPI, non-pharmaceutical intervention (social distancing, masking, etc.); vax, vaccination.

Of the 17 rounds, 3 were classified as Decision designs (decision x decision), 6 rounds as VOI designs (decision x uncertainty), and the other 8 as some form of uncertainty x uncertainty design (Table 1). The Decision designs were clustered earlier in the pandemic (December 2020-March 2021), representing a period when decisions regarding NPIs and vaccines were most needed. The VOI designs occurred throughout the pandemic (January 2021-April 2023) and focused on various vaccination decisions, such as increase of primary series coverage among adults (Round 2), expansion of the vaccine program among children (Round 9), or comparison of different booster strategies (Rounds 14 and 15). In these VOI designs, the second axes typically described properties of virus variants, extents of waning, or immune escape. Of the 8 rounds classified as uncertainty x uncertainty designs, 6 were considered as situational awareness, 1 as sensitivity analysis, and 1 as horizon scanning. Situational awareness rounds were designed to anticipate the arrival of new variants, or evaluate the potential impact of growing vaccine hesitancy and declining NPIs. The round classified as sensitivity analysis was devoted to understanding the impact of waning assumptions on disease dynamics (training Round 8 in summer 2021, which was not publicly released). The round classified as horizon scanning explored potential interactions between waning immunity and a hypothetical immune escape variant in the post-Omicron period. Although SMH scenarios span many of the designs presented in our proposed classification, forecasting scenarios were not represented per se, in part because the primary purpose of the SMH was not to explicitly combine scenarios (see Bay et al., *in review* for a post-hoc application of this concept).

These classifications were challenging to make because the SMH rounds were used (and implicitly designed for) many audiences, each of which might interpret a design differently. For example, Round 4 was designed specifically with the CDC in mind in Spring 2021, and, at the time, the degree to which they should emphasize vaccination versus compliance with NPIs was important (Decision category, decision x decision axes). A similar classification would apply from the lens of a state or county public health agency, because recommendations to the public about vaccination and NPIs were in their authority. In contrast, for a hospital administrator, these scenarios might have served as situational awareness (uncertainty x uncertainty) that could was useful in anticipating staff and resource needs over the coming months. Relatedly, Rounds 11 (December 2021) and 12 (January 2022) addressed the Omicron variant and were designed primarily for situational awareness, with scenarios informed by early data on variant characteristics from South Africa. However, given the limited amount of information available on Omicron severity in Round 11, a broad range of severity assumptions was chosen, so that this round could also be considered horizon scanning.

Another interesting challenge in retrospectively classifying the designs of the 17 SMH rounds was judging whether a particular axis was a decision or uncertainty axis, as the same axis designed for the same user might have had a different meaning at different stages of the pandemic. For instance, in the first four rounds, we interpreted the vaccination and NPI axes as decision axes, because the CDC and other public health agencies were actively grappling with how aggressively to recommend vaccination, how to allocate initially limited doses of vaccine, and how strongly to implement and enforce NPIs. In Round 5, however, by May of 2021, public health agencies seemed to have become somewhat resigned to the behavioral choices of individuals regarding vaccination and compliance with NPIs, and so we treated those factors as uncertainties rather than decisions. On reflection, it would have been easier to classify the axes and the scenario designs in the moment, and in consultation with the motivating audiences.

### Detailed Considerations in Scenario Design

In the following sections, we describe the elements of a scenario design in an epidemiological setting, more elements of experimental design that are pertinent, and other practical considerations. To guide the reader, we have provided a figure that recapitulates the process of scenario design and highlights key components that need to be considered (Figure 3). When relevant, we illustrate these considerations with examples drawn from the 17 rounds of SMH scenario designs.

**Figure 3.**
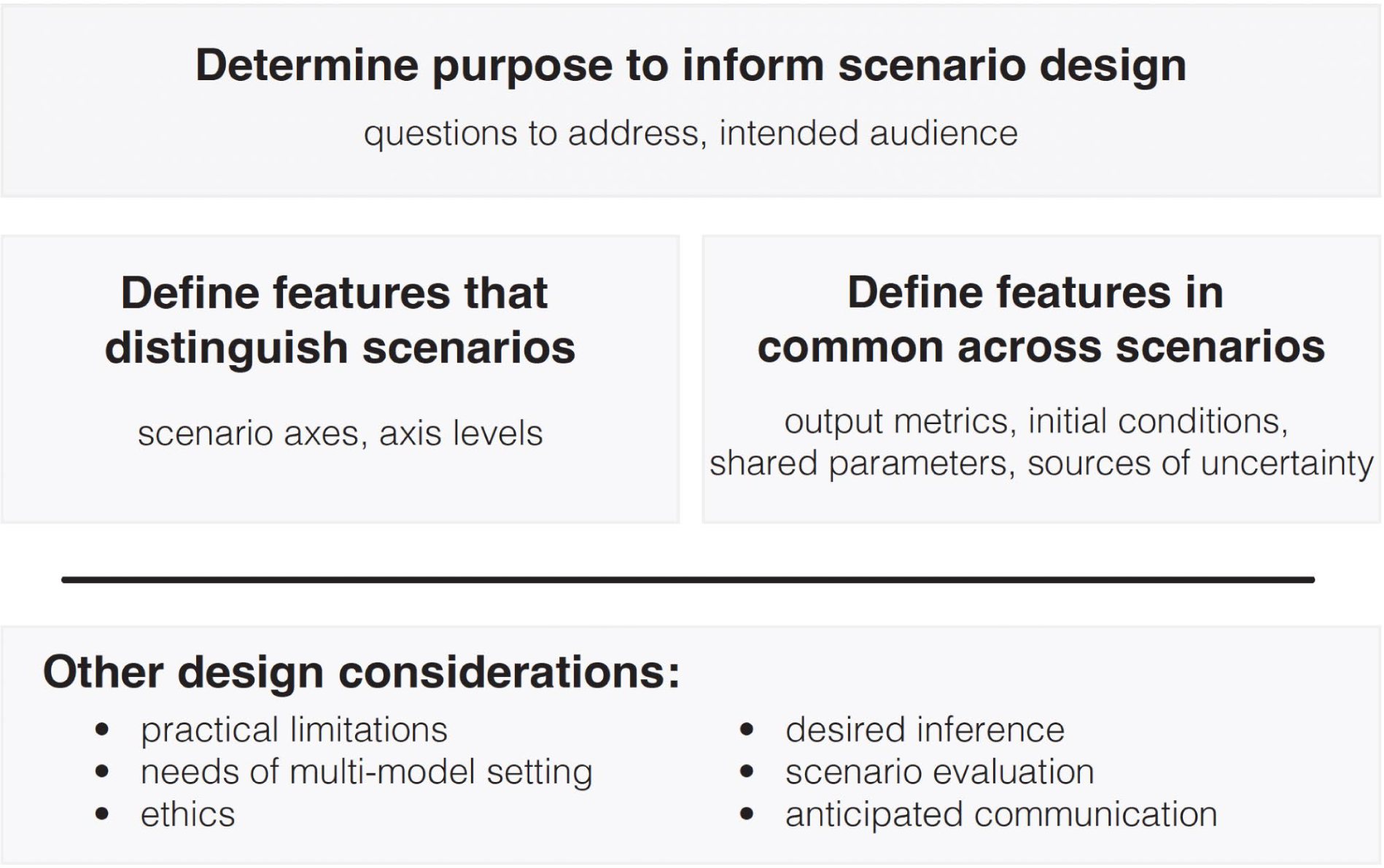
Overview of scenario design process. First, determine the purpose of the scenario modeling exercise, including the questions to be addressed and the intended audience. This purpose informs all other design decisions. The taxonomies defined in Figures 1 and 2 should be applied at this step. Then, define the features that distinguish scenarios and those that are common across scenarios. Last, consider other design issues that may be relevant during all phases of scenario design.

#### Elements of a Scenario Design

Several key elements should be considered in scenario design, including output metrics, details of how the scenarios differ, initial conditions, common factors, and the approach for handling uncertainty not otherwise expressed across scenarios. All such design decisions should be informed by the scenario’s purpose. Transparency and precision is important for communication, especially in multi-model settings to ensure that contributing models produce comparable outputs.

##### Output metrics

A scenario design needs to specify output metrics to be projected. This choice can strongly influence the insights gained as well as interventions recommended (Probert et al., 2016). For infectious disease projections, output metrics might include incident or cumulative cases, hospitalizations, or deaths, which may be further broken down by subgroups (e.g., age, region, race, or ethnicity). Metrics that are not strictly epidemiological may also be of interest (e.g., business closure duration for NPI-based scenarios, Shea et al., 2023). Generally, multiple outputs are assessed separately, but multi-criteria decision analysis (Keeney and Raiffa, 1993) would permit composite outputs to be addressed. The scenario design also needs to specify the time frame and spatial extent of outputs, including temporal and spatial resolution. Another important consideration is the establishment of shared “ground truth” data for output metrics, particularly in the context of a collaborative effort, as these data are typically used for model Details about how to summarize and report results also should be specified; this will depend in part on the desired inference. A full probability distribution for the output metrics might be desired, or summaries like the mean, median, or an exceedance probability might suffice. Alternatively, individual replicates (i.e., daily or weekly simulations) may be useful to contrast projections across scenarios (see discussion about pairing replicates in *Experimental Design* section). Individual replicates also offer more flexibility than summary measures, as the need for nuanced metrics may arise after scenarios have been designed. We refer the reader to Sherratt et al. (*in review*) for deeper comparison of summary outputs and individual replicates.

##### Distinguishing features of scenarios

The key aspects that differentiate the scenarios are the choice of axes and levels set for each (Fig. 3). Earlier, we discussed how to choose scenario axes; here, we provide guidance on setting levels.

Levels on a decision axis may represent specific discrete interventions (e.g., approve a vaccine for a specific age group or not) or represent a continuous variable that is closely tied to a decision (e.g., high and low coverage of a vaccination campaign). In the latter case, the levels chosen might bracket the range of effects that could be achieved under different strategies. Sometimes counterfactual scenarios are used to evaluate the population-level benefits of an intervention. For example, at the beginning of COVID-19 vaccine rollout in December 2020, SMH Round 1 considered a scenario without any vaccination as well as scenarios with various coverage levels (Table 1). Because the vaccine had already been approved and manufacturing was in progress (FDA, 2020), the no-vaccination scenario was not expected to eventuate, but it was important for comparative purposes.

Levels on an uncertainty axis may be based on available estimates of the parameter of interest and it is common to set values using the associated confidence intervals. For sensitivity analysis or situational awareness designs, values associated with an 80- to 95-percent confidence interval may represent reasonable bounds on current knowledge (Fig. 1B). Horizon scanning designs may use more extreme values to illustrate what could happen if the future does not conform with the past (Fig. 2B). When empirical confidence intervals are not available, expert opinion, literature review, or survey information can be used to bracket optimistic and pessimistic assumptions (e.g., SMH has used behavioral surveys of propensity to get vaccinated, Beleche et al., 2021). Further considerations on the choice of levels, as related to experimental design, are discussed below.

##### Common factors

Another important aspect of scenario design is the factors that are common across the scenarios. These commonalities are not part of the scenario axes and can include shared data sources and their interpretation, common assumptions about disease dynamics, behavioral responses, or interventions.

##### Initial conditions

Initial conditions represent the state of the modeled system at the start of the scenario projection period. The initial conditions may vary across scenarios or across models. For example, differing scenario assumptions about waning immunity may not only imply a different understanding of what will happen in the future, but also about what occurred in the past; thus, calibration of the model could lead to different initial conditions for each scenario. In a multi-model setting, precise initial conditions are rarely defined, because the models have different calibration approaches and structures. Instead, it is valuable to specify aspects of the process that all models should employ to set initial conditions.

##### Additional sources of uncertainty

While well calibrated forecasts integrate over all sources of uncertainty, scenarios typically encompass a subset of all possible uncertainties (Reich et al., 2022). Forecasting scenario designs represent a special case of scenarios, where the combination of uncertainty captured within a single scenario and between scenarios should be comprehensive (see Fig. 2C). For all other scenario designs, judgment can be made about how much uncertainty to include. The power to discern differences among the scenarios increases as other factors are controlled, but this comes at the expense of generalizability. In a collaborative hub setting, it is important for different modeling teams to make their own choices about many of the uncertainties not specified in the designs. However, it can sometimes be valuable to provide guidance for how to handle key parameters and assumptions that could drive disease dynamics that are not part of the scenario axes (e.g., all SMH rounds provided guidelines on vaccine efficacy, and bounds were often prescribed for waning immunity).

#### Principles of Experimental Design as Applied to Scenarios

As noted above, scenario design and experimental design are closely related conceptually and structurally. Individual scenarios are analogous to experimental treatments, and there are analogous considerations of replication, randomization, and control. Here we briefly discuss these parallels.

The scenario designs proposed in our taxonomy have analogs in experimental design. For example, a 2×2 VOI design is analogous to a randomized block design, where the uncertainty axis serves as a control (or block) variable, to test whether the intervention effect is consistent across blocks (Montgomery, 2020). Designs with more than two axes or levels per axis are also possible, and fractional factorial designs (where only a subset of the full factorial design is explored) can be used to explore the main effects of many factors, without having to run as many scenarios. Also, there is a tension in statistical design of experiments that helps choose the levels of the factors: the closer the levels are together, the more reasonable it is to assume a linear effect between them; but the farther the levels are apart, the higher the power to discern differences and the scope of inferences that can be made. Similar logic is applicable in scenario design.

In experimental design, holding all factors constant within each replicate is a powerful form of control. Pairing replicates across scenarios is an analogous concept, where as many elements of the model as possible are matched in a particular replicate, like the initial conditions, the sampled parameter values, and, if possible, some aspects of temporal variance. Although common in some fields (e.g., McGowan et al., 2011), pairing replicates can be challenging in epidemiological models, especially if the initial conditions depend on the scenario specification or if demographic stochasticity (e.g., binomial sampling for individual outcomes) is integral to the model (Kaminsky et al., 2019). Nevertheless, even if it is partial, pairing replicates increases the power to discern treatment effects.

In some cases, multiple rounds of experiments are anticipated, and the results from the early experiments can be used to refine later experiments. Similarly, sequential designs can be achieved with multiple rounds of scenarios. Several SMH rounds were sequential updates in response to the arrival of a new variant or to inform a new policy. For instance, Round 14 was designed to inform the ACIP recommendation for reformulated boosters in the fall of 2022, comparing age-restricted versus broader coverage. Presenting results to policymakers (Rosenblum et al., 2022) prompted a follow-up round (Round 15), which made small changes to scenario axes and values to assess whether there would be benefits to releasing boosters earlier (Table 1). If a comparison of outputs across sequential rounds is planned, it is important to record factors that may change between rounds, confounding outputs of interest (e.g., changes in data availability, types of interventions being considered, or new model developments).

More complex scenario designs are possible with multi-round scenarios, including dynamic sequential and branching scenarios. Sequential designs are not fixed *a priori,* but depend on the outcomes of experiments during the exercise (Wald, 1947; Robbins, 1952; Chernoff, 1992). Methods to analyze such sequential statistical designs can be employed to analyze sequential scenarios. Branching scenarios, motivated by branching or nested statistical designs (Hung et al., 2009), can be combined with sequential elements to produce scenarios that are valuable for training and tabletop exercises, where the branches arise in response to dynamic interventions made by users (Barrett et al., 2015; Parikh et al. 2016).

#### Other Design Considerations

##### Practical limitations

There are practical limitations and trade-offs in scenario design, including model capability, computational resources, clarity of assumptions, and time taken to design a scenario that is actionable. Scenarios must not be too complex, so that modeling teams can generate projections in a reasonable amount of time. In a multi-model setting, minimally complex scenarios also encourage participation from a larger number of teams. Access to additional computational resources can be enhanced in times of crisis, but the need to balance the aims of the scenario design with the practical aspects of modeling remains. If practical constraints strongly and repeatedly influence scenario design, the purposes of the scenario may need to be revisited.

To accommodate these multifaceted needs, the scenario design process is often iterative, involving both internal and external discussions. Internal communication of scenario requirements, especially in the context of a multi-model hub, usually requires multiple rounds of discussion to reduce unwanted (linguistic) uncertainties while retaining a good expression of the scientific uncertainties focal to the scenarios (Shea et al., 2020). If time permits, something akin to the modified Delphi process is valuable: produce a first round of results; discuss the results across models as a group, looking for differences arising from linguistic uncertainty; then allow teams to produce a second round of results that reflect the clarifications (Shea et al., 2020). However, if decision makers only have a short period of time to implement an intervention, a small number of simple scenarios run on stripped-down models might be all that can be achieved.

External discussions with public health decision makers can inform the choice of scenario axes (e.g., potential interventions) and corresponding assumptions (e.g., compliance with those interventions). Curiously, evidence from cognitive psychology suggests that decision makers often need help to fully articulate their concerns (Bond et al., 2008), so a back-and-forth conversation to develop the purpose is an important step. As a result, scenario design can sometimes take several weeks, as illustrated by the SMH experience (Table 1).

##### Ethical considerations

Ethics of scenario design inherit attributes from the broader ethics of biomedical research (Beauchamp and Childress, 2009), epidemiological research (CIOMS 2009), and decision-making for public health emergencies (Emanuel et al., 2022). Scenario design should have the properties of autonomy, beneficence, non-maleficence, and justice. Autonomy requires the scenario design process to be scientifically grounded and well-documented (including specification of clear objectives, Smith et al., 2021), capturing uncertainty and recording assumptions. Additionally, scenario design should be beneficial, in that it should promote evidence-based policymaking (Choi et al., 2005), providing benefit over decisions that would be made without scenario projections (Taylor, 2003). These principles also play a role in non-maleficence, as inaccuracies in scenario design may cause harm to populations affected by the recommendations. Existing inequalities should be incorporated into and addressed by scenario design; further, scenario design should not exacerbate these inequalities, nor create new inequalities (CIOMS 2009). Scenario modeling efforts should be evaluated according to these criteria (Boden et al., 2017). Multiple metrics of equity and fairness can be considered in scenario design (Braveman and Gruskin, 2003; Whitehead, 1992; Mhasawade et al., 2021), as different stakeholders may have different perspectives (Whitehead, 1992).

##### Scenario evaluation

In some cases, there may be a desire to evaluate scenarios and projections after the projection period has passed; does this desire affect scenario design? Broadly, scenarios are well designed if the resulting projections answer the primary question and serve the intended users even if the scenario assumptions do not materialize. Yet it still may be useful to assess how well scenario assumptions match unfolding reality, especially when a goal of scenario design is bracketing (i.e., situational awareness or forecasting situations). Scenario evaluation is difficult in practice; Howerton et al. (*in review*) provide an illustration of salient issues. Scenario parameters may not be measurable even after projection periods have passed (e.g., degree of immune escape of a new variant, or even the impact of an NPI). For horizon scanning and for scenarios including counterfactuals, evaluation of scenario parameters and resulting projections can be particularly difficult. Overall, being able to evaluate scenarios is not a requirement and does not need to be prioritized over other goals, although it is good practice where possible and can build trust with end-users.

##### Communication

In epidemiology, scenario projections are often designed for specific audiences and can have public visibility. Thus, it can be advantageous for scenarios to be clear enough for easy communication to and interpretation by external audiences. For example, SMH uses a standard scenario design template to provide consistency in how scenario assumptions are shared and ease comparisons between rounds and between hubs (see https://github.com/midas-network/covid19-scenario-modeling-hub for an example). For additional discussion of generalizable infrastructure, see the HubVerse project (https://hubdocs.readthedocs.io/).

Presentation of scenario results is also an important component of communication worth anticipating. A successful display of results entails three, often conflicting, objectives: enabling comparisons among scenarios; communicating the uncertainty within and across models (in a multi-model setting); and supporting multiple different classes of constituents, including researchers, public health officials, journalists, and members of the public. Key challenges include communication of the nuances of scenario projections to lay audiences (as different from forecasts), and visualization of uncertainty (Kamal et al., 2021; Hullman et al., 2019; Hägele et al., 2022; Spiegelhalter, 2017). See Loo et al., *in review* for further discussion of SMH communication strategies.

## Discussion

The experience of the U.S. COVID-19 Scenario Modeling Hub over its first 17 rounds provided an impetus for the scenario taxonomy proposed in this work, which we believe will be valuable in epidemiological settings, and perhaps more broadly. We have attempted to provide broad guidelines for scenario design that apply in single and multi-model efforts, and made parallels with other fields such as experimental design. Several insights with broader relevance bear reflection: the importance of the audience and a clear statement of the purpose of the design; the power of the design itself; the need to think carefully about uncertainty; and the benefits of a clear process.

By projecting multiple, clearly defined scenarios that were motivated by public health needs, SMH projections have had significant public health impact (Borchering et al., 2021, 2023; Truelove et al., 2022; Rosenblum et al., 2022; Biggerstaff et al., 2022). It was difficult at times, however, to balance the needs of decision makers with the capabilities of available models. Implementing realistic scenarios and generating well-calibrated projections can require added model complexity or additional time. Key policy questions or vast uncertainty may suggest the need for many scenarios, but computational constraints may limit the number of scenarios that can be modeled in a timely fashion. Further, the foundational philosophy behind multi-model ensembles, namely, the diversity of approaches taken by the independent groups (Shea et al., 2020) can itself pose a challenge for scenario design. But the repeated nature of the SMH effort has allowed the complexity of the models and the subtleties of scenarios to increase.

Clarity of audience and purpose affect scenario design and its impact. The influence of SMH rounds that were developed in direct conversation with a decision-making agency was easiest to illustrate. But many valuable impacts are harder to demonstrate, like the deepening understanding among modeling teams of the epidemic in the U.S. prompted by the structured challenge of shared scenarios. In retrospect, we found it somewhat challenging to look back over two years of work and recover the specific purposes of each round. We propose that an active and clear articulation of the audiences and the purposes of a scenario design will help to sharpen the design of the scenarios in future SMH rounds, and similar efforts in the future.

The design of scenarios provides the structure for inference. The taxonomy captured in Figures 1 and 2 was not available during the design of SMH Rounds 1 through 17, but we believe that it could have enhanced some of the designs. Value of information designs are particularly interesting, because they both allow the comparison of alternative interventions and test those comparisons against critical sources of uncertainty. The SMH used VOI designs in 6 of the first 17 rounds, but curiously, none of those showed a reversal in preference of intervention based on the uncertainty axis. On one hand, that’s a great relief to decision makers, but on the other hand, it raises a question about whether the uncertainties considered were most relevant to the decisions. Would conscious attention to the power of particular design structures lead to even more valuable scenario designs?

Scenario design invites careful and deliberate consideration of uncertainties. Scenario axes often focus on uncertainties hypothesized to be major drivers of future dynamics or decision outcomes. The first question is whether the process used to identify those uncertainties is robust. The second, perhaps more difficult question, is how to handle the remaining uncertainty. For example, operational uncertainties about the implementation of interventions may be required to create clear, easy-to-interpret scenarios, but such uncertainties are also important to account for in projected outcomes. In evaluating outcomes of the early SMH rounds, Howerton et al. (*in review*) and Wade-Malone et al. (*in review*) note that results of individual models often had quite different variances, suggesting that they captured different sets of uncertainty that weren’t otherwise specified in the scenario design. Is that problematic or desirable? How does calibration of the individual models affect calibration of the ensemble projection (Howerton et al., 2023), and how does that affect scenario design? We believe that there are some open questions here that warrant further study.

The process of scenario design affects efficiency, participation, trust, and communication. Particularly in multi-model collaborative settings, the process of scenario design is challenging, and a clear process with dedicated support staff can support and invite the participation of the collaborating teams. But even in single-model settings, the process of scenario design aids in communication with the intended audiences and can promote trust.

As noted earlier, scenario design is practiced in many fields besides epidemiology. The framework that we have proposed in this paper integrates elements of the three schools described by Amer et al. (2013): like the Intuitive Logics School, it relies on experts’ conceptual understanding of systems to develop causal maps that inform scenario design; like the Probabilistic Modified Trends School, it combines extrapolation of past trends with modifications to acknowledge changes in the future; and, like the French School, it places an emphasis on the decision setting, that is, the ways in which trajectories can be influenced by intervention. We are hopeful that more cross-disciplinary examination of how scenario projections are designed and used can lead to common advances across fields.

Beyond the insights that arose from the SMH experience, there are other questions that may be relevant for development of scenario modeling practices in epidemiology. We see these as open questions for a future research agenda, to improve the impact of scenario methods:

- Are there perspectives in scenario design, especially in collaborative ensemble settings in other fields, that would enhance the practice in epidemiology? Similarly, can our efforts inform practice in other fields?
- Is there a design trajectory across an epidemic? That is, can we anticipate a specific sequence of questions, and even have template scenario designs ready? As a proposal, four stages could arise: (1) initial bounding of uncertainty and exploration of simple interventions; (2) assessing specific interventions (e.g., vaccination) as they become available; (3) assessing new dynamics (e.g., variants, behavior changes) as they arise; and (4) transitions to questions relevant in an endemic phase. Is this a useful start?
- What are the pitfalls to avoid in scenario design in public health settings? Is it possible to inadvertently mislead or confuse decision makers with poorly designed scenarios? Are other unintentional negative outcomes possible?
- Were there an operational scenario modeling hub for a particular disease, would a set of scenario designs become standard? For instance, would a regularly calibrated baseline scenario with several updated contrasts (e.g., emergence of a new variant) make sense? Or are infectious diseases too complex, because human behavior, available interventions, and viral evolution change so quickly that standard scenario designs are not useful?
- Can lessons from scenario design in one location or outbreak reliably be applied in other settings? This will be particularly important at times where urgent results are needed, or in low-resource settings (e.g., low and middle income countries).
- How can we best communicate scenario results and explain the difference between scenarios and forecasts, which are more intuitive? Are verbal or numerical or graphical representations of scenario designs and results most effective and do they differ for more or less quantitatively comfortable users?

In summary, the COVID-19 pandemic coalesced a great deal of burgeoning expertise in epidemiological modeling, scenario projection, scenario design, and collaborative modeling endeavors. Using the experience of the U.S. Scenario Modeling Hub to reflect on the state-of-the-art in scenario design, we believe that a sound philosophical framework and procedural methodology for scenario design would increase the efficiency and efficacy of these methods, both in epidemiological settings and in other fields of endeavor.

## Data Availability

All data produced in the present work are contained in the manuscript

## Acknowledgments

We are grateful to all the participants in the U.S. COVID-19 Scenario Modeling Hub, for stimulating conversations over the last two-and-a-half years that have motivated the ideas in this paper. KS, EH, and RB acknowledge funding from NSF COVID-19 RAPID awards DEB-2028301, DEB-2037885, DEB-2126278 and DEB-2220903. EH was supported by the Eberly College of Science Barbara McClintock Science Achievement Graduate Scholarship in Biology at the Pennsylvania State University. KY was supported by the Penn State Verne M. Willaman Distinguished Fellowship in Science and NSF Grant No. DGE1255832. HH was supported by NIGMS Grant U24GM132013. MM, SV, and BL acknowledge support from NSF Expeditions in Computing Grant CCF-1918656 and VDH Contract UVABIO610-GY23.

## Disclaimers

The findings and conclusions in this report are those of the authors and do not necessarily represent the official position of the US National Institutes of Health or Department of Health and Human Services. Any use of trade, firm, or product names is for descriptive purposes only and does not imply endorsement by the U.S. Government.

## Declarations of Interest

MCR reports stock ownership in Becton Dickinson & Co., which manufactures medical equipment used in COVID-19 testing, vaccination, and treatment. JL has served as an expert witness on cases where the likely length of the pandemic was of issue. There are no other competing interests to declare.

